# Seroprevalence and attainment of herd immunity against SARS CoV-2: A modelling study

**DOI:** 10.1101/2021.01.22.21250328

**Authors:** Abhijit Paul, Harshith B Kadnur, Animesh Ray, Samrat Chatterjee, Naveet Wig

## Abstract

**Objective:** The present study is aims to predict the likelihood of and likely time required to attain herd immunity against COVID-19 in New Delhi due to natural infection.

**Method:** An ODE based mathematical model was constructed by extending the classical SEIR model to predict the seroprevalence rate in Delhi. We estimated the parameter values for Delhi using available data (reported cases and the seroprevalence rate) and used them for future prediction. We also attempted to capture the changes in the seroprevalence rate with different possibilities of reinfection.

**Results:** Maximum seroprevalence rate obtained through our model is 31.65% and also a reduction in the seroprevalence rate was observed for the upcoming one month (month of January, 2021) due to the reduced transmission rate. After increasing the transmission rate to the value same as the third wave in New Delhi, we obtained a maximum value of 54.96%. This maximum value significantly decreased with the reduction in the reinfection possibilities. Also, a little impact of the duration of persistence of antibodies, 180 vs 105 days, was observed on the maximum seroprevalence.

**Conclusion:** This modelling study suggests that natural infection alone, as gauged by serial sero-surveys, will not result in attainment of herd immunity in the state of Delhi.

## Introduction

COVID-19, first reported from China on 16th December 2019, has spread globally affecting more than 91 million people causing 1.9 million deaths. Indian scenario is following the global trends with 10 million cases and 151,918 deaths (as of 16th January, 2021) [1]. The pandemic is still raging on in different parts of the world showing periodical waxing and waning.

Immune response against SARS CoV-2 is still not completely defined. Antibodies test (IgG against SARS CoV-2), being a surrogate method of measuring immune response, has been utilized in a number of studies to detect past infection with COVID-19. Studies have shown that the immune response is not uniform among all the infected individuals and only ∼80% of the asymptomatic individuals mount a measurable antibody response [2,3]. Also IgG antibody titres start reducing within 2-3 months post COVID-19 infection [4]. Longevity of immune response against COVID-19 is yet to be entirely defined and reinfections are already very well documented [5].

Oxford dictionary defines herd immunity as “resistance to the spread of an infectious disease within a population that is based on pre-existing immunity of a high proportion of individuals as a result of previous infection or vaccination”. Herd immunity threshold for COVID-19 required is estimated to be 60% to 75% based on the equation (1-1/*R*_0_) [6, 7]. Few policy makers and groups of scientists around the world proposed allowing unmitigated spread of infection to attain herd immunity by natural infection. Relaxing social restrictions at the peak of pandemic at various places has been suggested to be part of this strategy. However, WHO and other health authorities, on the other hand, have warned against targeting the strategy of attaining herd immunity by natural infection as it entails, potentially significant mortality and morbidity, especially among high risk individuals [8].

Seroprevalence surveys are being conducted in various cities to quantify spread of COVID-19 infection as well as to inform various policy decisions. Monthly sero-surveys have been conducted in New Delhi between July and October. However, the trend of seroprevalence in these studies did not increase substantially over several months[9]. With documented rapidly declining antibody response and reported reinfection, will we be ever able to reach the threshold required for herd immunity? To answer this question, we constructed a mathematical model to predict the time required to attain herd immunity against COVID-19 in New Delhi.

## Materials and methods

### Data Acquisition

The state-wise data was collected from an open-sourced database for COVID-19 stats & patient tracing in India [10]. In this study, we have considered the day-wise confirmed, recovered and deceased cases in Delhi till January 16, 2021. Seroprevalence data between July and October available for New Delhi were used [9].

### Mathematical model

We propose an extended epidemic model (see **Figure 1**) which incorporates four infectious classes: pre-symptomatic infectious (*P*), undetected infectious (*U*), tested infectious (*T*), reported infectious (*R*_*I*_). A large portion of infectious individuals do not show any symptoms or show mild symptoms [11], thus a portion of such infectious individuals remain undetected [12,13]. Tested infectious (*T*) individuals are those who went for testing but results are not yet known. After knowing the result, the individuals with COVID-19 positivity will enter the reported infectious (*R*_*I*_) class. These reported infectious (*R*_*I*_) individuals usually do not transmit the disease due to home isolation or hospitalization. The susceptible individuals (*S*) acquire infection from pre-symptomatic infectious (*P*), undetected infectious (*U*) and tested infectious (*T*) individuals and enter into exposed class (*E*). It is to be mentioned here that an undetected infectious (*U*) individual may enter into the tested infectious (*T*) class by rapid antigen or other tests like Reverse transcription polymerase chain reaction (RT-PCR) test. We also incorporated two scaling parameter *α*_*U*_ and *α*_*P*_ for undetected and pre-symptomatic infectious class, respectively, to make the infection spreading rates different from the tested infected ones. Transformation from *U* and *R*_*I*_ classes to undetected recovered (*U*_*R*_) and recorded recovered (*R*_*R*_) classes occur through the rates 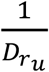 and 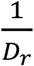, respectively. The death rates due to this disease are denoted by 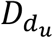 and *D*_*d*_ for these both *U* and *R*_*I*_ classes respectively, and these individuals join into the undetected death (*U*_*D*_) and recorded death (*R*_*D*_) classes. Here, the total population(*N*) is assumed to be constant, which is a reasonable assumption if the epidemic period is not too long. Though re-infections are increasingly being reported in COVID-19, the knowledge about exact frequency, risk factors and period of susceptibility is still evolving. So, we have introduced a parameter (*σ*) in our model to incorporate the different possibilities of reinfection. The dynamics of these different classes across time (t) are described by the following set of differential equations:

**Figure 1:**
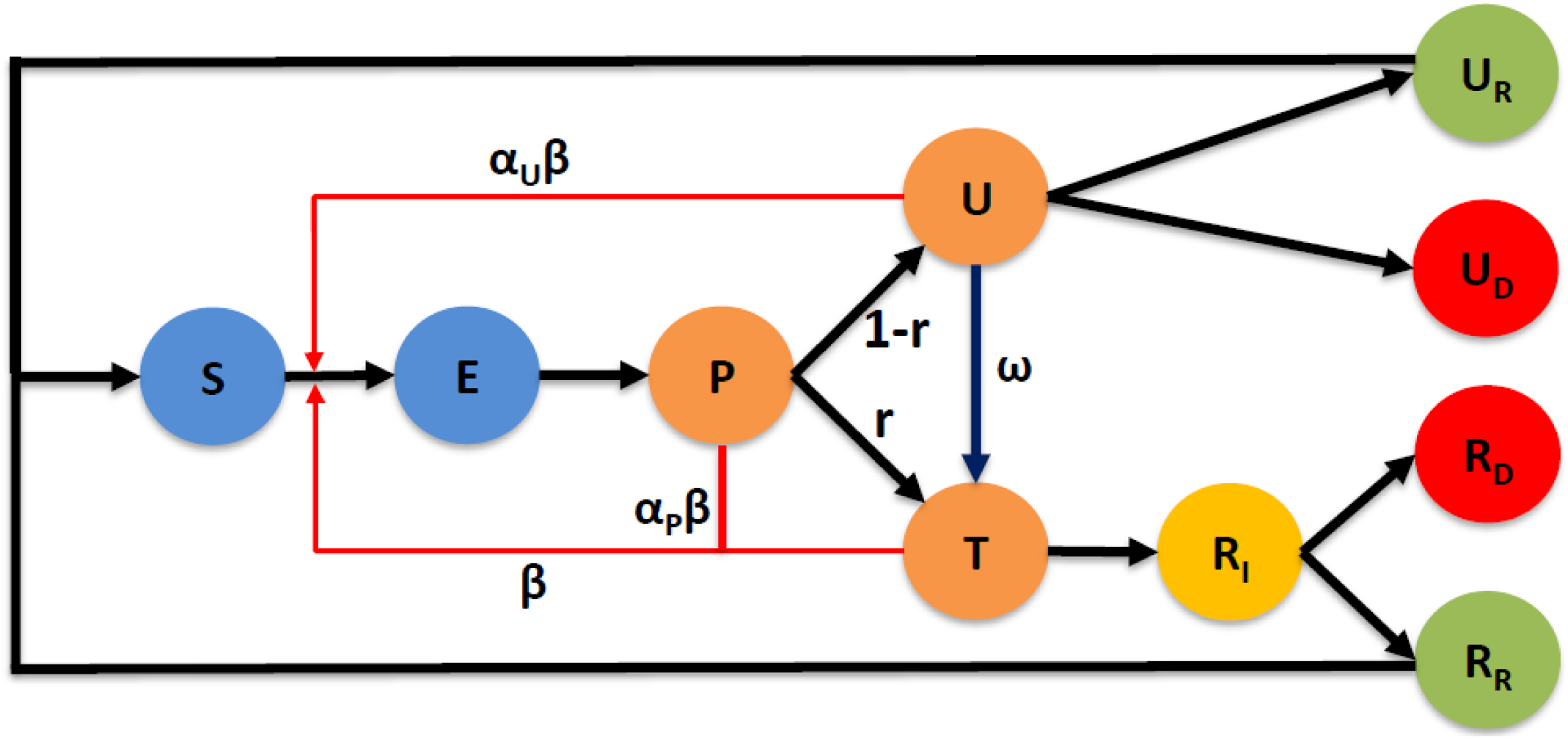
Schematic diagram of the proposed model. We extended the classical epidemic model (SEIR) by incorporating ten compartments: susceptible (*S*), exposed (*E*), pre-symptomatic infectious (*P*), undetected infectious (*U*), tested infectious (*T*), reported infectious (*R*_*I*_), undetected recovered (*U*_*R*_), undetected death (*U*_*D*_), recorded recovered (*R*_*R*_) and recorded death (*R*_*D*_). We have also included the case where the recovered individuals become susceptible again due to loss of antibodies.

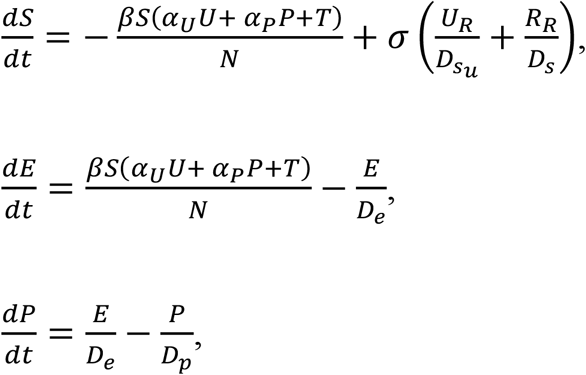

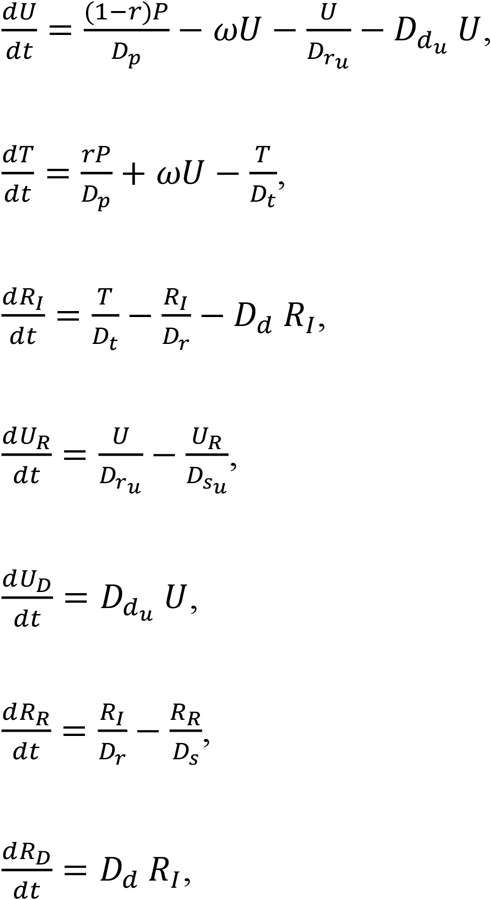

with *S* + *E* + *P* + *U* + *T* + *R*_*I*_ + *U*_*R*_ + *U*_*D*_ + *R*_*R*_+ *R*_*D*_ = *N* (total population) & *S*(0), *E*(0), *P*(0), *U*(0), *T*(0), *R*_*I*_(0), *U*_*R*_(0), *U*_*D*_(0), *R*_*R*_(0), *R*_*D*_(0) ≥ 0. Definitions and values of the parameters with reference are provided in **Table 1**. Seroprevalence rate at a given time (t) are obtained from the following equation:

**Table 1:**
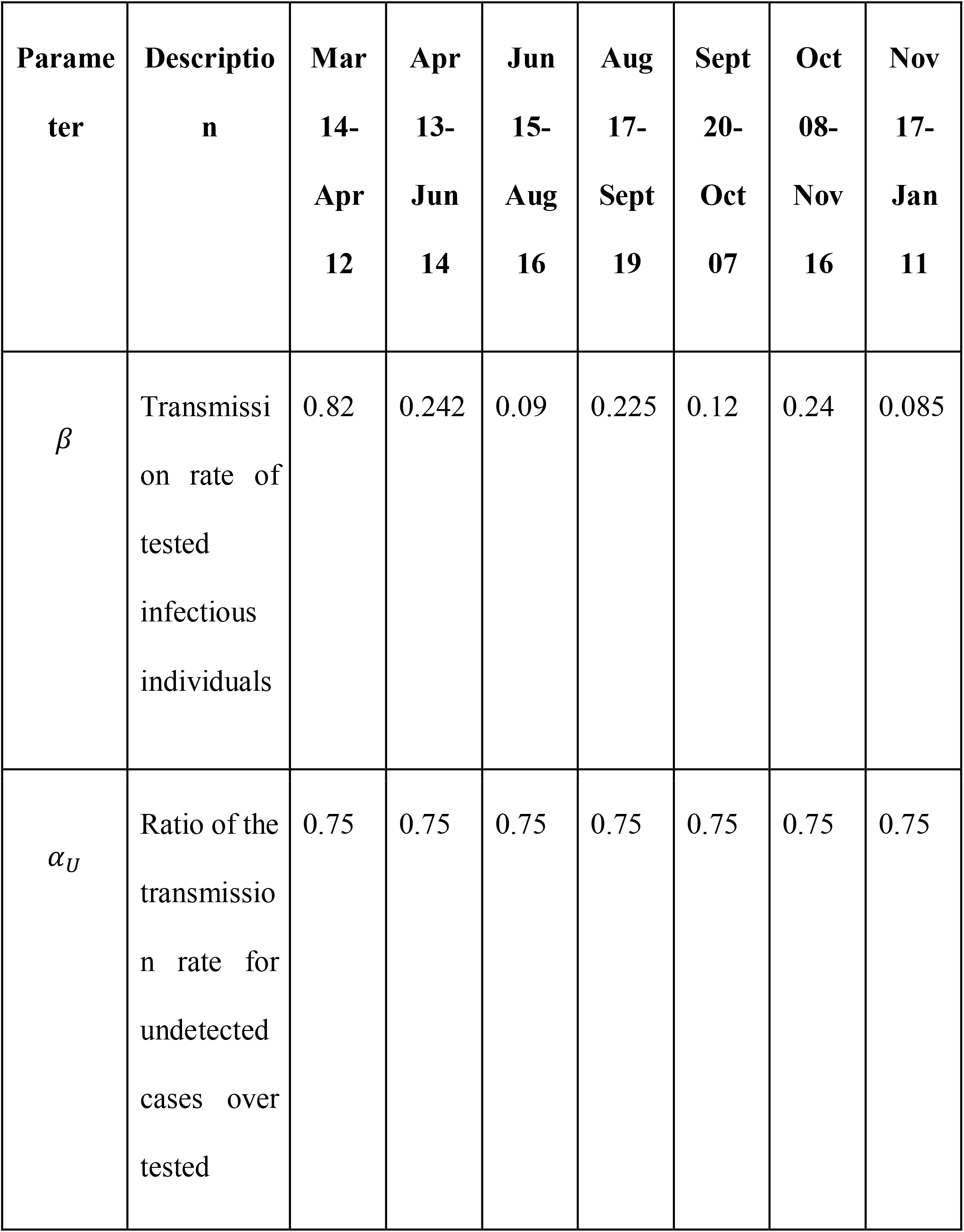
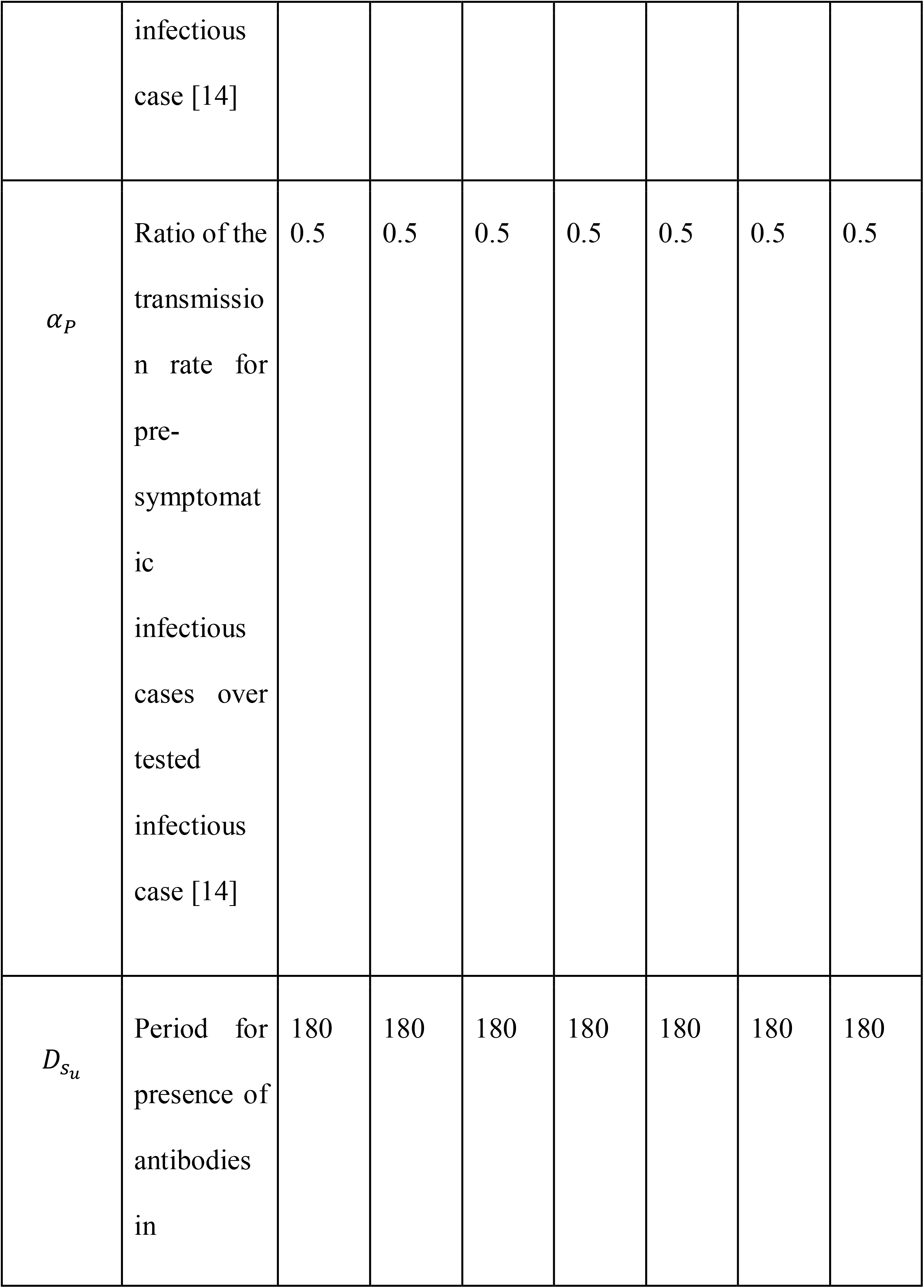
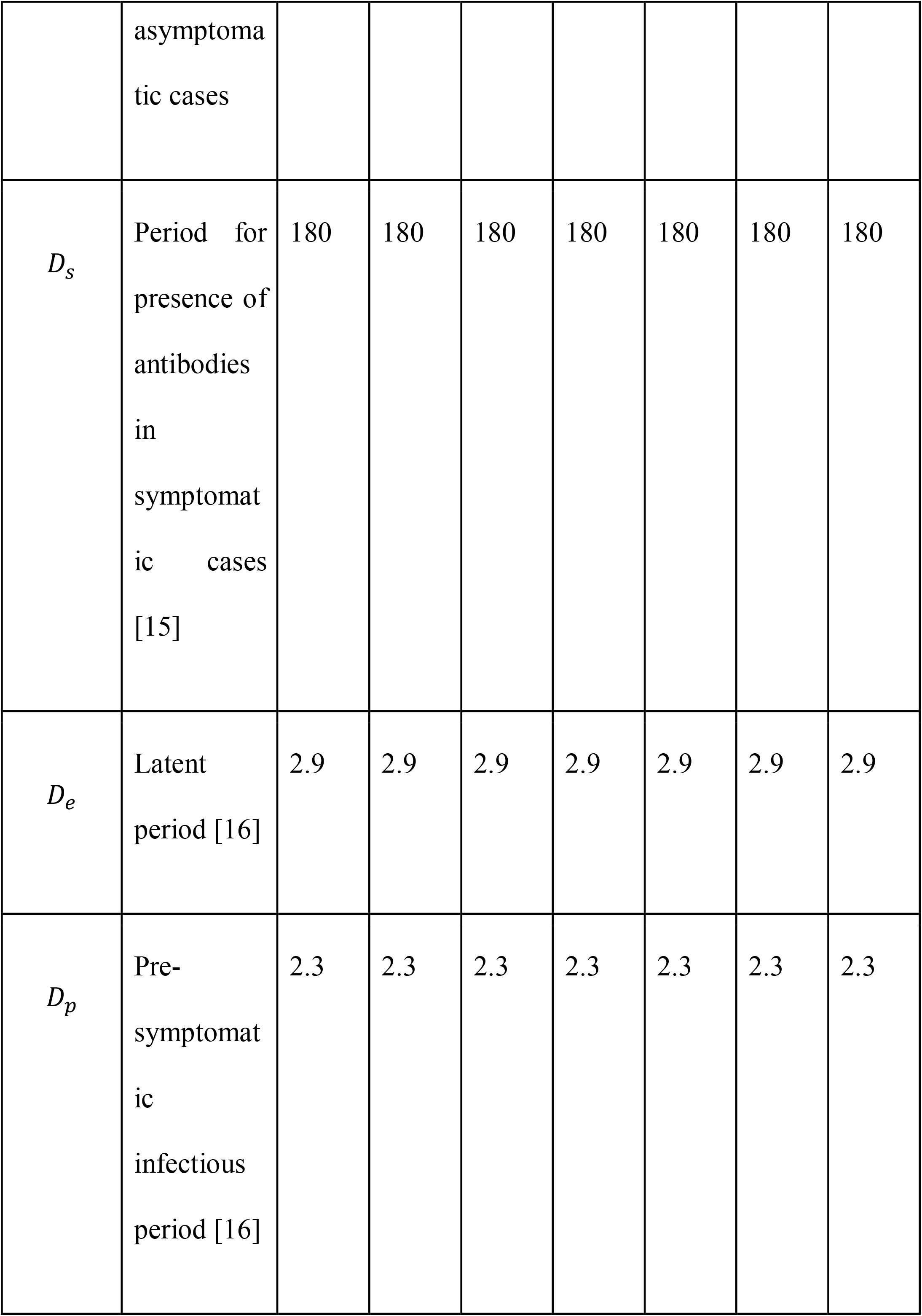
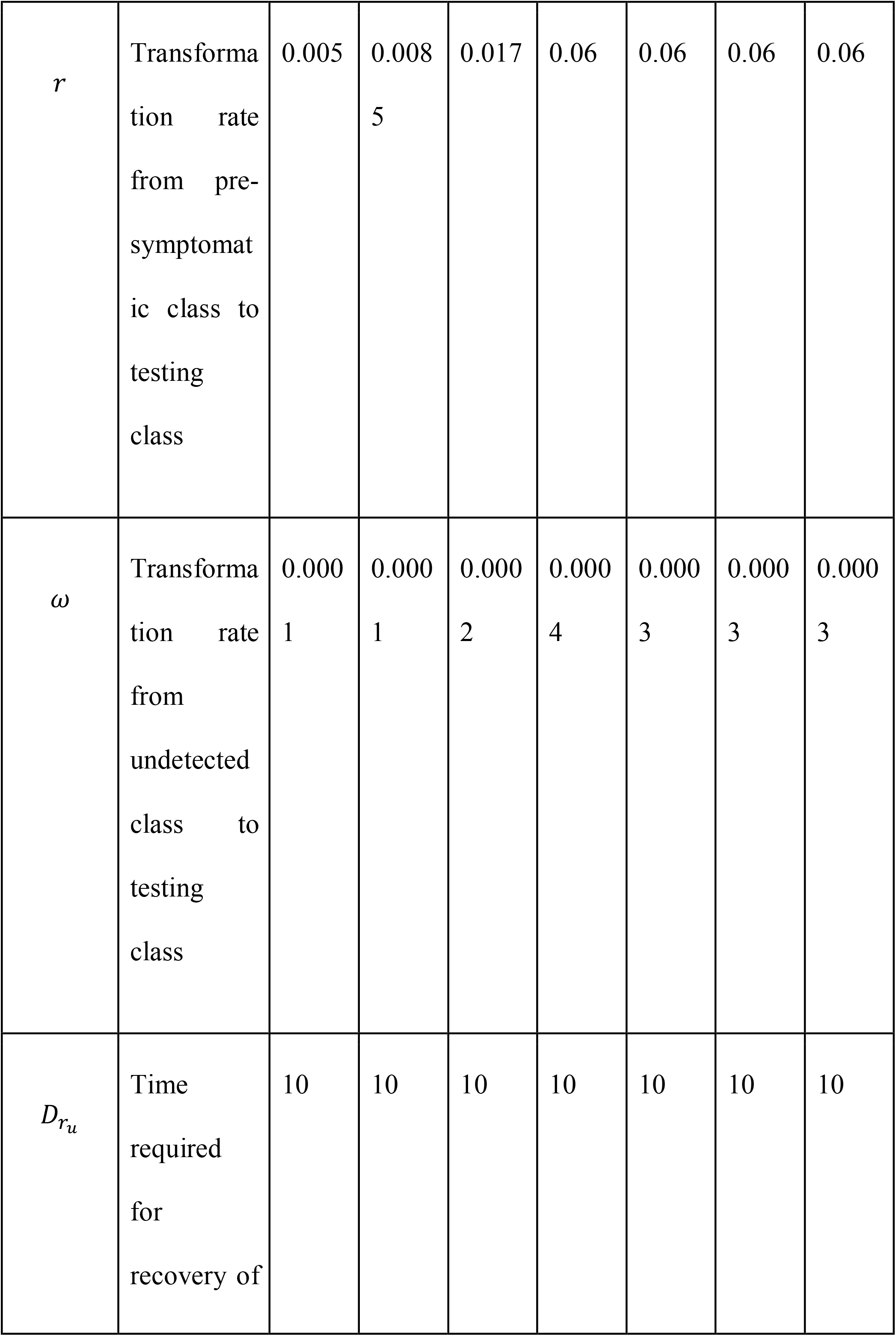
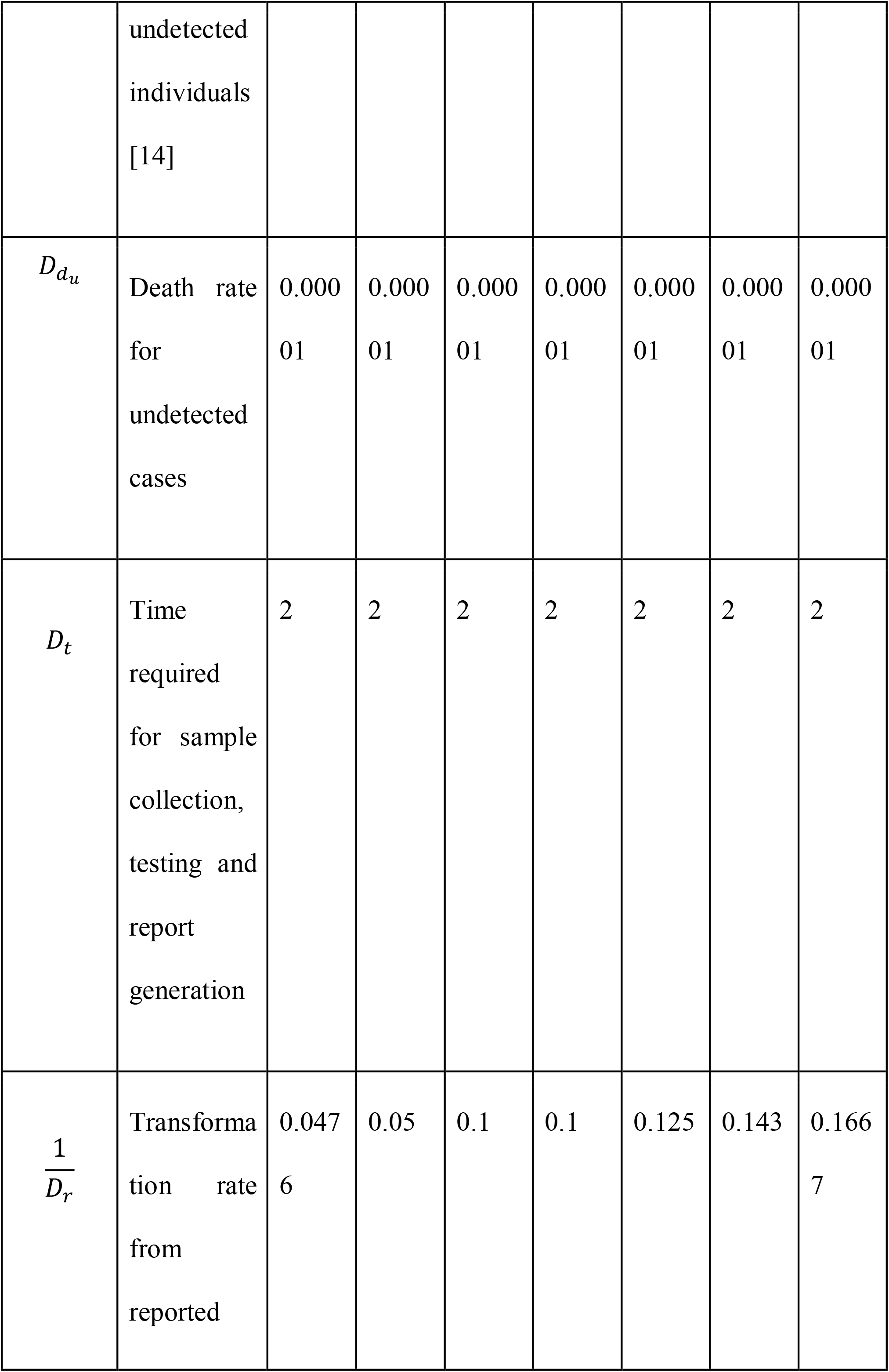
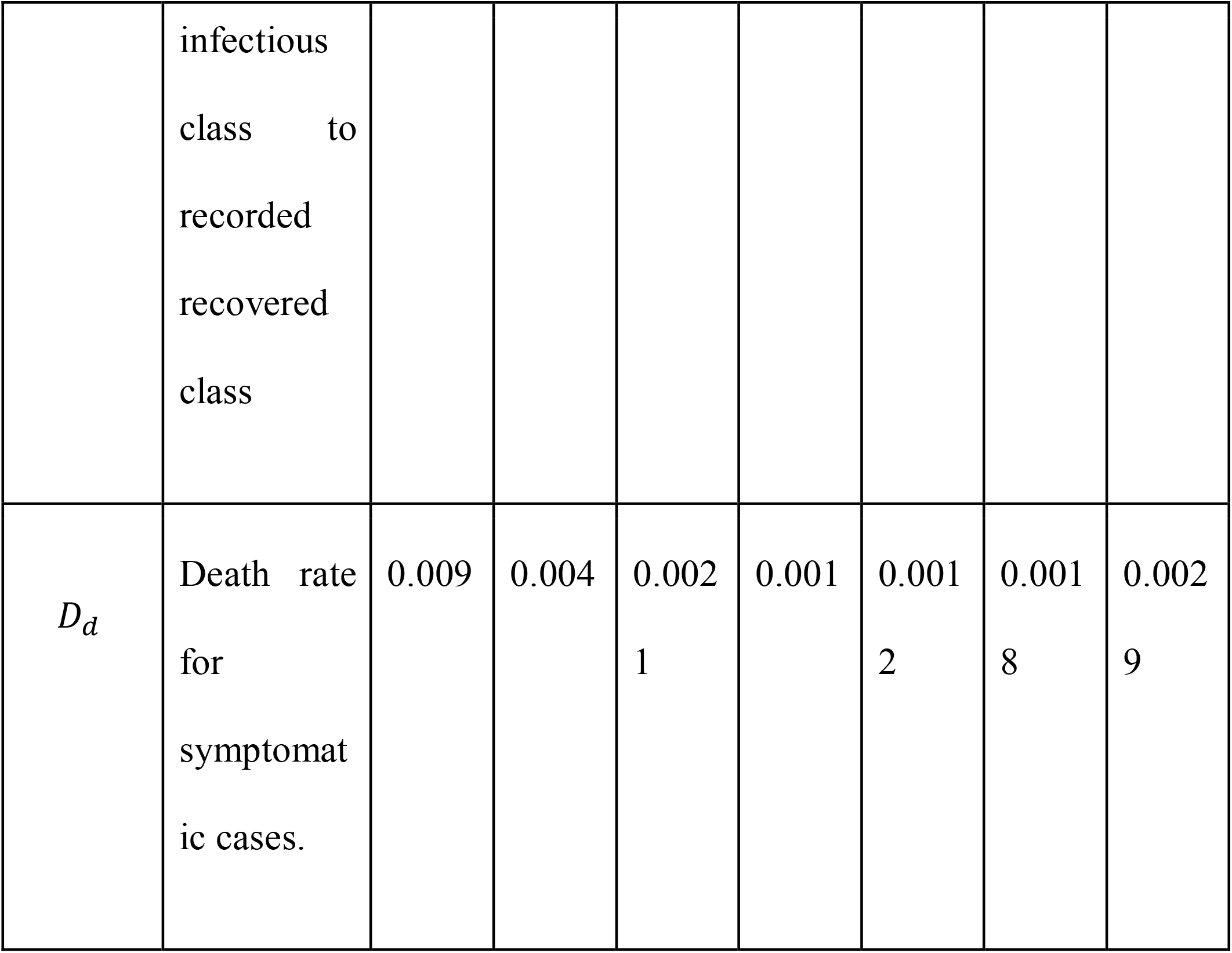
Parameter setting for the model simulation. Parameters for which the references were obtained are provided in the description section and remaining were estimated according to good fit.

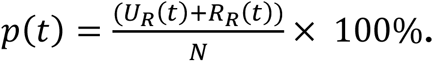

### Numerical simulation

All the numerical simulation for this mathematical model was performed in MATLAB by using the ODE45 function. Also, the curve fitting toolbox was used to get the R-squared value between the simulated and predicted data. Among the parameters considered in modelling, a few required estimation by trial and error method to best fit the curve **(Table 1)**.

Prediction of the number of confirmed, active, recovered and deceased cases for the upcoming one month were done by considering the parameter set obtained for the period of Nov 17, 2020-Jan 16, 2021 range. This allowed us to maintain the changes in the number of confirmed, active, recovered and deceased cases with the same rate obtained at the end of the study period.

## Results

### Model Analysis

The aim of this study was to capture and predict the future seroprevalence rate in Delhi through mathematical modelling. We extended the classical SEIR model by including pre-symptomatic infectious (*P*), undetected infectious (*U*) and reported infectious (*R*_*I*_) classes (see methods and **Figure 1**) and considered the daily data of confirmed, active, recovered and deceased cases in Delhi till January 16, 2021 [10]. The model estimated time-series curves for confirmed, active, recovered and deceased cases are provided in **Figure 2** along with their reported data. The obtained R-squared value of >0.97 indicates a good fit of the model results. The parameter values corresponding to time-series curves are presented in **Table 1**, which was further used for predictions.

**Figure 2:**
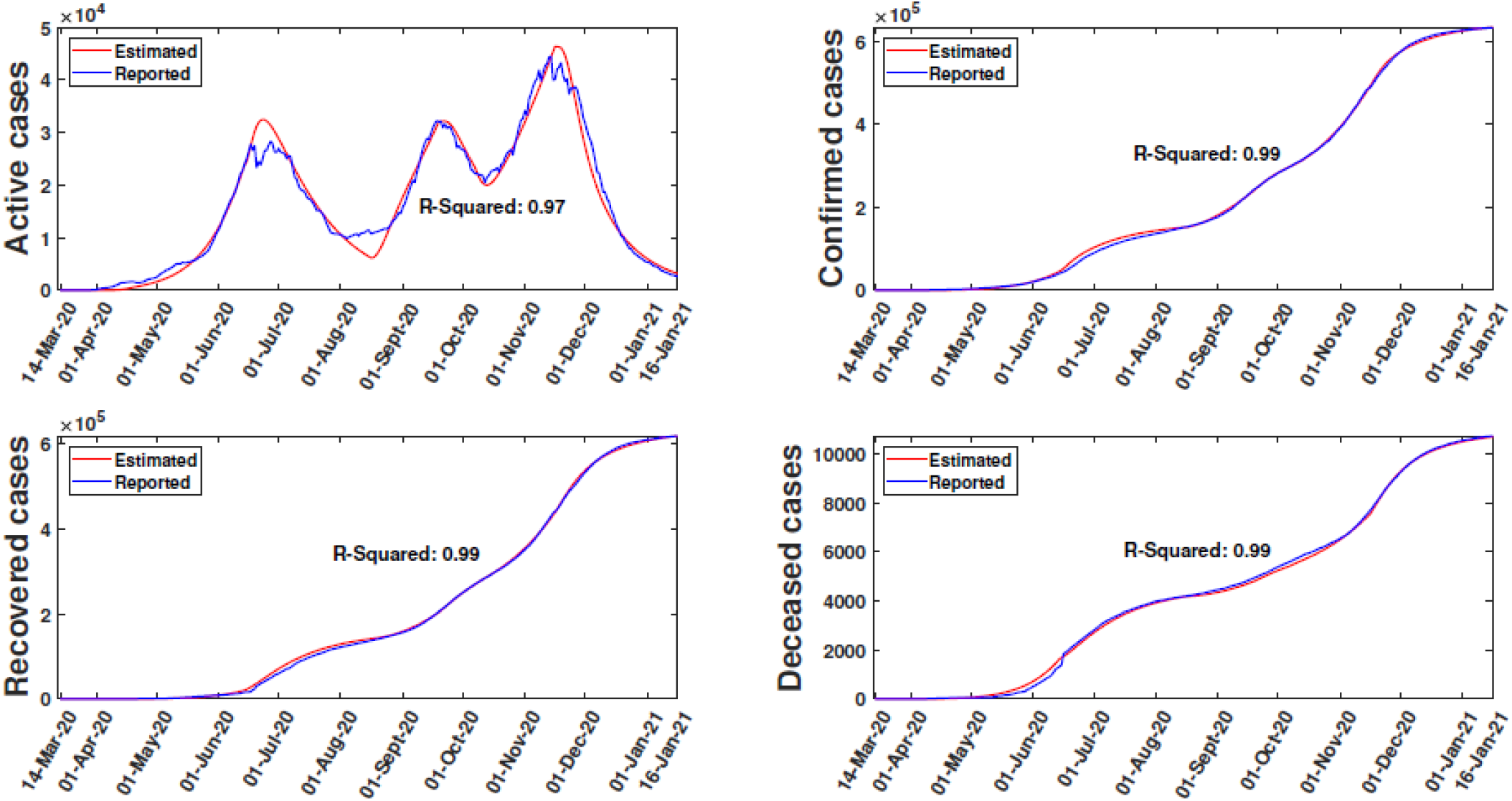
Comparison between the reported data (blue colour curve) and the model simulation data (red colour curve) of Delhi. The data contains day-wise information for the period March 14, 2020 to January 16, 2021 for Delhi and the parameter values are given in **Table 1**. The R-squared values are also mentioned here in the inset.

### Model Prediction

We used the parameter set obtained for the period of November 17, 2020 to January 16, 2021 range (see **Table 1**) to predict the number of confirmed, active, recovered and deceased cases for the upcoming one month (month of January, 2021) (**Figure 3**). We also plotted the variation in the seroprevalence rate with respect to time (**Figure 4**) for the whole study period. Here, we were able to capture the reported seroprevalence rate of New Delhi for the month of July, September and October but not for August [9]. We observed that from around the last week of November this value reached above 30% and from the 2 ^nd^ week of December, the seroprevalence rate started to fall. During our study period, we obtained a maximum seroprevalence rate around 31.65 % on December, 8. Also, a reduction in seroprevalence rate was observed for the upcoming one month (month of January, 2021) due to the reduced transmission rate in the period of Nov 17, 2020-Jan 16, 2021.

**Figure 3:**
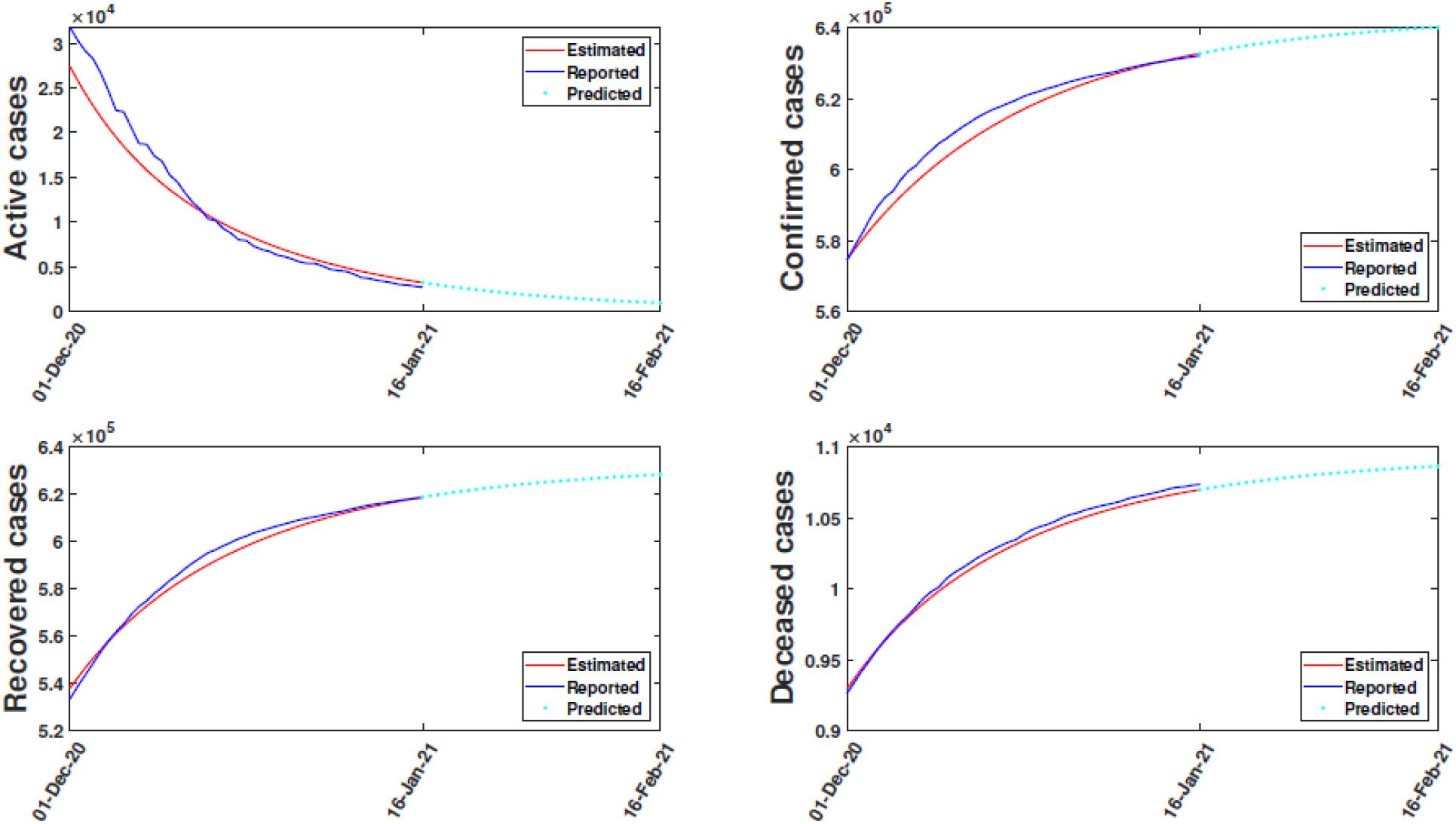
Number of confirmed, active, recovered and deceased cases after one month from January 16, 2021. We have used the parameter set of the period of Nov 17, 2020-Jan 16, 2021 range (see **Table 1)** for this prediction.

**Figure 4:**
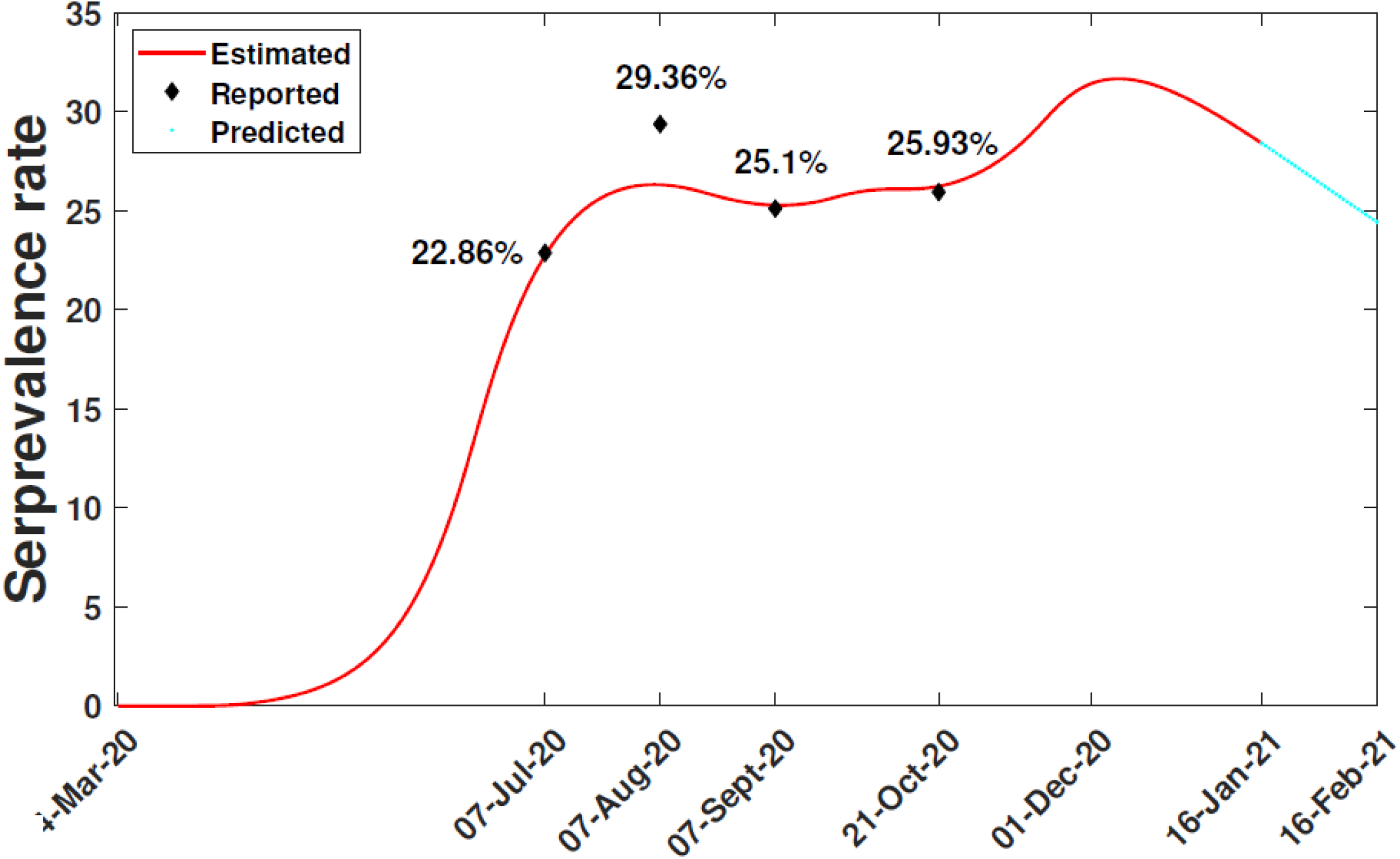
Variation in estimated seroprevalence rate with time. The reported seroprevalence rates for July 1 ^st^ week, August 1 ^st^ week, September 1 ^st^ week and October 3 ^rd^ week are also provided as diamond shapes. In the last week of November this value is expected to reach above 30%. Thereafter, it is predicted to fall and reach around 24.4% at the end of the study period.

We tried various simulations to understand the possibility of getting herd immunity by achieving a > 65% seroprevalence rate, as calculated by the formula = 1-1/*R*_0_[18]. Thus, we increased the value of transmission rate to 0.24, a value same as that during the third wave (Oct 08, 2020-Nov 16, 2020). We observed that the seroprevalence rate was projected to go beyond 50% after 156 days before falling, after reaching the maximum value of 54.96% (see **Table 2** and **Supplementary Figure S1**). In this case, we have considered 100% reinfection possibility of the individuals who lose their anti-SARS CoV-2 antibodies. Next, we attempted to capture the changes in the maximum seroprevalence rate with the different reinfection possibilities. Thus, we considered 95%, 75%, 50% and 25% reinfection possibilities and found that the maximum seroprevalence rate decreases with the reduction in the reinfection percentage (see **Table 2**). We also changed the persistence of antibodies from 180 days to 105 days in these five different reinfection possibilities and found significant reduction in the maximum seroprevalence rates (see **Table 2** and **Supplementary Figure S1**).

**Table 2:** Predicted maximum seroprevalence rates for different reinfection possibilities. Here, we considered an increased value of transmission rate with the same obtained for the third wave for each case. The variation of the seroprevalence rates with time for these different cases are provided in **Supplementary Figure S1**.

| **Reinfection possibility** | **Persistence of antibodies (in days)** | **Maximum seroprevalence rate** | **Time point** |
| --- | --- | --- | --- |
| 100% | 180 | 54.96% | 196 days after January 16, 2021 |
|  | 105 | 51.86% | 165 days after January 16, 2021 |
| 95% | 180 | 51.58% | 203 days after January 16, 2021 |
|  | 105 | 49.47% | 164 days after January 16, 2021 |
| 75% | 180 | 40.3% | 209 days after<br>January 16, 2021 |
|  | 105 | 35% | 173 days after<br>January 16, 2021 |
| 50% | 180 | 31.77% | 201 days after<br>January 16, 2021 |
|  | 105 | 32.4% | December 3,<br>2020 |
| 25% | 180 | 31.43% | December 9,<br>2020 |
|  | 105 | 32.65% | December 4,<br>2020 |

## Discussion

In our study, we saw that the maximum seroprevalence achievable in Delhi was less than 65%, suggesting that herd immunity, as predicted by serial sero-surveys, may not be achieved by natural infection alone. The maximum seroprevalence achievable was 54.96%, assuming that the re-infection rate and transmission rate was very high. Maximum seroprevalence achievable when parameters were unaltered was much lower (only 31.65%). The duration of persistence of antibodies, 180 vs 105 days, had minimal impact on the seroprevalence rate according to the prediction by our model. In both the scenarios, the required threshold of herd immunity i.e. 65% was not achieved.

Various factors leading to failed herd immunity could be,

1. Antibody response to COVID-19 though robust starts fading off in the early convalescent phase [2]. Immunity against SARS CoV-2, measured as antibody response, lasts for 3-6 months rendering the individuals susceptible to reinfection, which though rare is well documented [5].
2. Secondly, among the asymptomatic individuals, only ∼80% mount a measurable antibody response. While those that do not develop antibodies, potentially are susceptible to reinfection.

The most important implication of our study are

1. If we consider seroprevalence to be surrogate of immunity against SARS CoV-2 in the community, natural infection alone cannot lead to attainment of herd immunity.
2. Repeated sero-surveys may not be much informative, at least till the time a sizable portion of the population is immunized.

To the best of our knowledge, this is the first study which has used modelling for predicting future trajectories of sero-surveillance and its implications on herd immunity. Interestingly, a study had suggested the presence of possible cross reacting antibodies for explaining the trend of sero-survey result in New Delhi and had predicted the seroprevalence rate in the city for the month of September [17]. However, unlike our ODE based dynamical model it was an entirely statistical model. A good internal validation score makes our study a near-accurate prediction model. Also various levels of re-infections and duration of persistence of antibodies were considered to gauge the impact on seroprevalence.

Interestingly, sero-survey of Mumbai showed decrease in seroprevalence from 40% to 33.4% in the second survey, similar to that of New Delhi [19] which suggests that a similar trend in Mumbai may be expected, as predicted for New Delhi. Results of serial sero-survey in other cities are not available and hence difficult to extrapolate.

Nevertheless, some limitations noted in our model are,

1. We have considered data of only one state (New Delhi) which may not be extrapolated to other states or regions. Since, average seroprevalence for the whole state was taken, population heterogeneity in the different districts or zones were not accounted for.
2. In this model, antibody response, as measured by sero-survey, was taken as surrogate for immunity. Role of CMI (cell mediated immunity), which may have persisted after waning of IgG antibodies, has not been studied in sero-surveys, and was not considered in the above model.
3. The 29% mark of seroprevalence of August 2020, could not be accounted for in our study.
4. Though our findings were validated internally, the availability of sero-surveillance data for only 4 months made external validation improbable.
5. Intervention with vaccines will have an impact on transmission rate, which was not considered in the prediction model.

## Conclusion

This modelling study suggests that natural infection alone, as gauged by serial sero-surveys, will not result in attainment of herd immunity in the state of Delhi, highlighting the importance of continuing COVID appropriate behaviour and role of widespread implementation of vaccination against COVID-19.

## Data Availability

No data is included as it is a modelling study

## Notes

### Competing Interest Statement

The authors have declared no competing interest.

### Funding Statement

No funding has been received for this modelling study

### Author Declarations

Appropriate research reporting guidelines have been followed

### Summary of Updates

Author list updated

