## Supplementary figure 1 for "Seroprevalence and attainment of herd immunity against SARS CoV-2: A modelling study"

**Supplementary Figure S1: Variation in predicted seroprevalence rate for different reinfection possibilities**. Here, we considered two different values of duration of persistence of antibodies in the recovered individuals: (A) 180 days and (B) 105 days.

**Supplementary Figure 1**

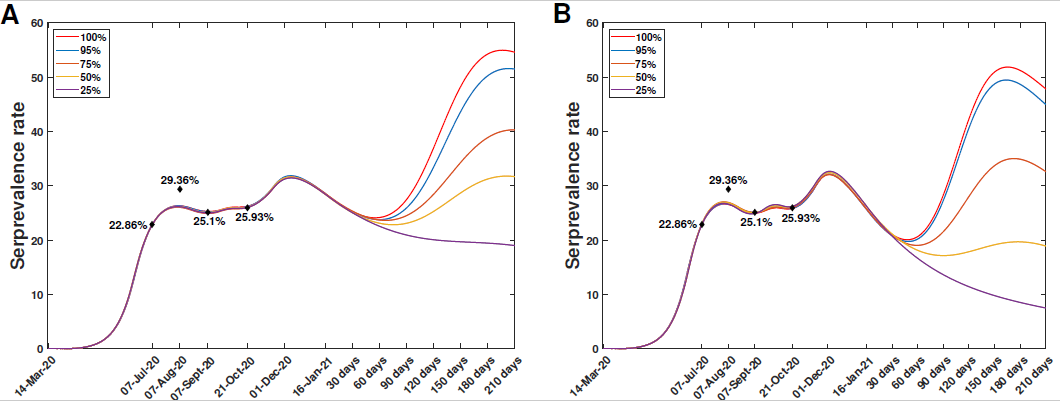
